# Single-Dose SARS-CoV-2 Vaccination With BNT162b2 and AZD1222 Induce Disparate Th1 Responses and IgA Production

**DOI:** 10.1101/2021.09.17.21263726

**Authors:** Michael Müller, Johann Volzke, Behnam Subin, Silke Müller, Martina Sombetzki, Emil C. Reisinger, Brigitte Müller-Hilke

## Abstract

While vaccination programs against SARS-CoV-2 are globally ongoing, disparate strategies for the deployment of spike antigen show varying effectiveness. In order to explore this phenomenon, we sought to compare the early immune responses against AZD1222 and BNT162b2. SARS-CoV-2 seronegative participants received a single dose of either vaccine and were analyzed for immune cell, effector T cell and antibody dynamics. AZD1222 induced transient leukopenia and major changes among innate and adaptive subpopulations. Both vaccines induced spike protein specific effector T cells which were dominated by Th1 responses following AZD1222 vaccination. A significant reduction of anti-inflammatory T cells upon re-stimulation was also restricted to AZD1222 vaccinees. While IgM and IgG were the dominant isotypes elicited by AZD1222, BNT162b2 led to a significant production of IgG and IgA. Our results suggest that the strategy for spike antigen delivery impacts on how and to what extent immune priming against the main SARS-CoV-2 antigen proceeds.

## 1 Introduction

The highly transmissible severe acute respiratory syndrome coronavirus 2 (SARS-CoV-2), which initially emerged in December 2019, has led to an unprecedented pandemic that caused over 4 Million casualties^1,2^. The prevailing occurrence of coronavirus disease 2019 (COVID-19) and its dramatic hazard for global health and economy has since spiked the rapid development of several vaccines. These collectively aim at the production of antibodies that will neutralize the binding of the viral spike glycoprotein to its angiotensin converting enzyme 2 (ACE2) receptor and thereby prevent cellular entry and subsequent infection^3–5^.

The urgent need to develop safe and efficient vaccines led to the deployment of various strategies, some of which were well established and others, like adenoviral vectors or mRNA, were novel. Among the early vaccines authorized by the European Medicines Agency (EMA) were the first-generation adenoviral vector AZD1222 that utilizes the simian dsDNA adenovirus ChAdOx1 as a vector for antigen delivery^6^. The first vaccine authorized by EMA was the nucleic acid based BNT162b2, a spike protein encoding N^1^- methyl-pseudouridine (m1Ψ) nucleoside-modified mRNA enveloped by lipid nanoparticles^7,8^.

Complete vaccination with either of the vaccines, which includes two doses at varying intervals, was shown to efficiently protect from symptomatic COVID-19^9,10^. Although early data hint at similar efficiencies after a single dose of either vaccine, booster immunization with BNT162b2 achieved somewhat higher rates of thwarting viral breakthrough^9–13^. With the emergence of SARS-CoV-2 variants that accumulate mutations in the spike glycoprotein^14–16^, the discrepancies between both vaccines grew even more pronounced with BNT162b2 leading to superior protection against the 1.351 (β) and 1.617.2 (δ)variants^15,17–19^.

We were curious about the molecular and cellular immune modules capable of mediating superior neutralization of SARS-CoV-2 and therefore aimed at exploring the immediate immune dynamics after a single dose of either AZD1222 or BNT162b2. To that extent, we investigated the proportions of peripheral leukocytes among innate and adaptive compartments over the first three weeks after immunization. To investigate the adaptive immune response in more detail, we surveyed the development of spike-protein specific plasma immunoglobulins as well as the re-activation and cytokine production of spike-specific effector T cells.

## 2 Results

### 2.1 Immune responses to AZD1222 and BNT162b2 differ quantitatively and qualitatively

A total of 40 participants were recruited from the local coordination center for clinical studies. Twenty of these participants were vaccinated with AZD1222 (Vaxzevria / Astrazeneca) and BNT162b2 (Comirnaty / Biontech), respectively. Two participants from each group had to be excluded retrospectively. One individual from the AZD1222 group because this subject was tested positive for antibodies at baseline and the others withdrew their consent for unknown reasons. Blood samples were obtained by venipuncture on the day of vaccination (day 0) and two, six, thirteen and twenty days later. Among all participants, 28 were available for all five consecutive venipunctures, five for four, two for three and one for two venipunctures. Table 1 lists the demographic data of all participants, showing an even distribution of sex and comparable age ranges between both vaccination groups.

**Table 1.** Demographics of study participants

|  | AZD1222 (n = 18) | BNT162b2 (n = 18) | p-value |
| --- | --- | --- | --- |
| sex [male/female] | 9 / 9 | 6 / 12 | 0.5236* |
| age [mean ± SD] | 36.7 ± 11.8 | 39.2 ± 11.5 | 0.4998<sup>#</sup> |

\*resulting from Fisher's exact test, <sup>#</sup>resulting from unpaired t test

In order to delineate the early immune cell responses to both vaccines, we performed 24-dimensional flow cytometry at each time point. Remarkably, by examining major immune cell compositions, we found a significant reduction (2.2-fold) in peripheral leukocytes on day 2 after vaccination with AZD1222 (Fig. 1A). This leukopenia resulted from significant reductions in granulocytes, B-lymphocytes as well as CD4- and CD8-positive T cells (Supplementary Table 1). When compared to baseline, leukocyte counts were still slightly reduced on day 6 yet back to normal on days 13 and 20. In contrast, vaccination with BNT162b2 did not result in any significant fluctuations among immune cell quantities (Supplementary Table 2).

**Fig. 1.**
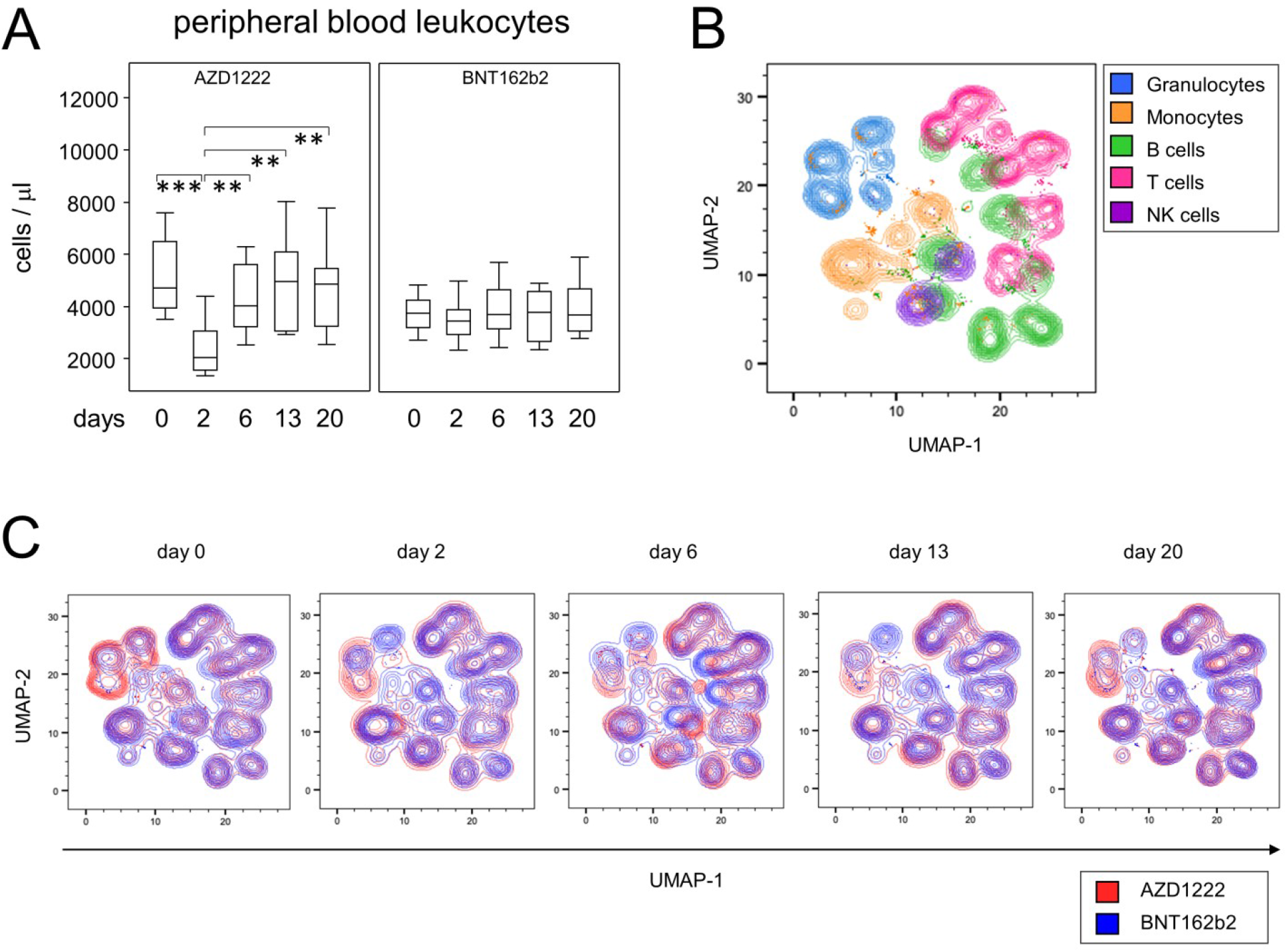
Vaccination with AZD1222 induced a transient reduction of peripheral leukocytes and displacements of major immune cell populations. **A** Leukocyte counts in the peripheral blood after vaccination with AZD1222 (n = 18, left panel) and BNT162b2 (n = 18, right panel). p-values resulting from multiple group comparisons were < 0.0001 (AZD1222 / Kruskal Wallis and Dunn’s multiple comparisons tests) and 0.6601 (BNT162b2 / one-way ANOVA), respectively. **p < 0.01, ***p < 0.001 **B** UMAP of surface antigen expression and clustering of major immune cell populations for all time points after vaccination. **C** UMAPs on immune cell compositions for each time point.

In order to survey qualitative alterations in the immune responses to either vaccine, we performed dimension reductions on our multiparametric data set by using the embedding algorithm UMAP. Fig. 1B summarizes all data from all time points and shows the topological distribution of immune cell subpopulations based on surface antigen expression patterns. Upon uncompressing the various time points, the overlay of both vaccine responses illustrates ample variation for the abundance of granulocyte, monocyte and B cell subpopulations primarily after administration of AZD1222, while differences regarding T- and NK cell subpopulations were less prominent for both vaccination regimens (Fig. 1C). Taken together, our data show that vaccination of SARS-CoV-2 seronegative participants with AZD1222 – unlike BNT162b2 – resulted in a transient reduction of peripheral leukocytes and led to alterations in immune cell compositions.

### 2.2 AZD1222 vaccination led to significant alterations among innate immune cell proportions

To further substantiate the time lines of early immune events following AZD1222 and BNT162b2 vaccination, we analyzed the major immune cell populations in more detail. E.g., live monocytes were identified by their sideward scatter properties before alterations of CD14 and CD16 expression patterns were analyzed for the various time points. Fig. 2A illustrates the time line for one participant receiving AZD1222. Fig. 2B documents a transient yet statistically significant increase in CD14^+^CD16^+^ pro-inflammatory monocytes on day 2 for the AZD1222 group (p < 0.0001). In contrast, there was no alteration among the proportions of pro-inflammatory monocytes following vaccination with BNT162b2 (Fig. 2B).

**Fig. 2.**
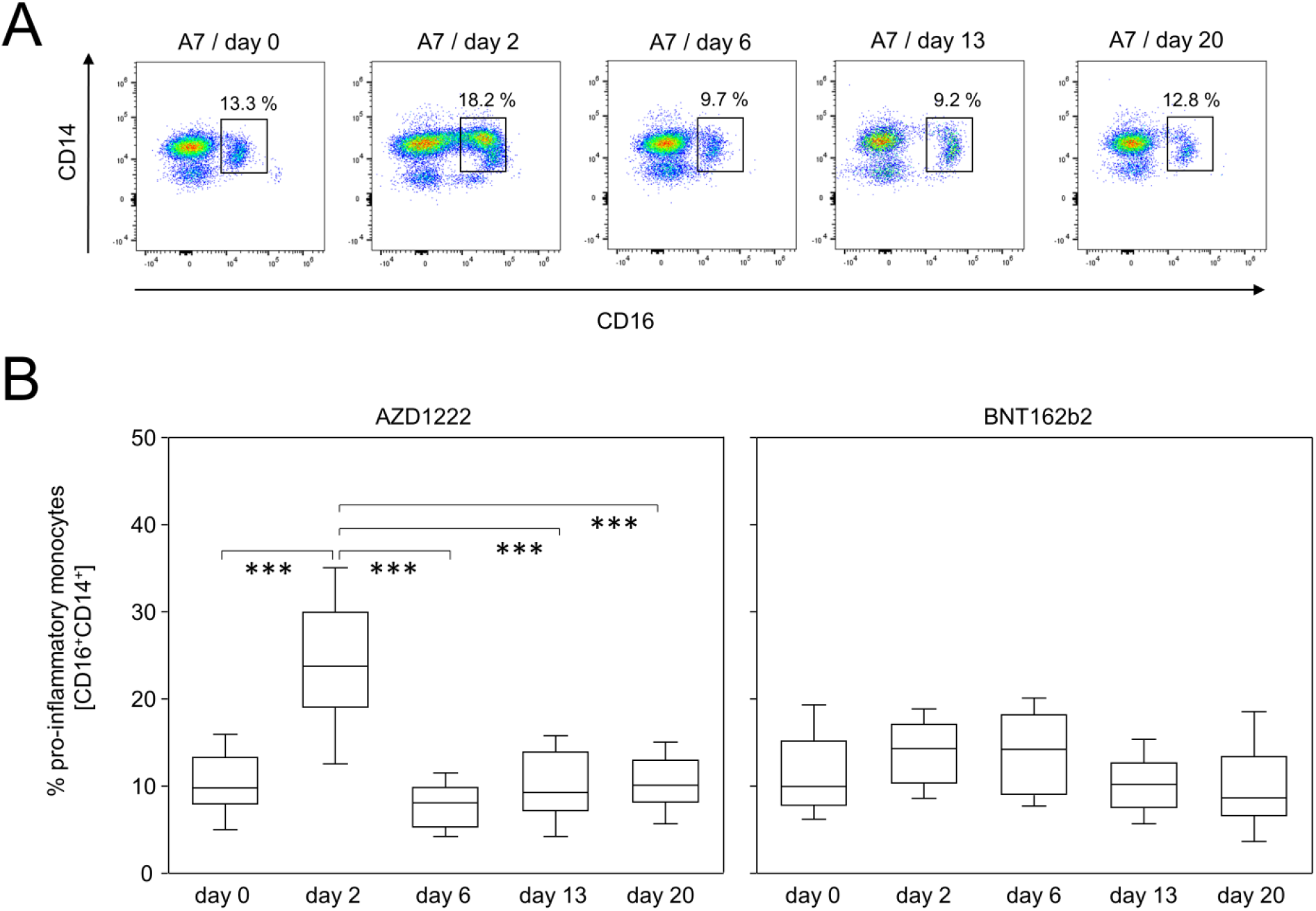
AZD1222 vaccination induced the enrichment of pro-inflammatory monocytes. **A** Pseudocolor plots for the expression of CD14 and CD16 on monocytes are representative for the AZD1222 vaccination group. **B** Proportions of CD16^+^CD14^+^ pro-inflammatory monocytes after vaccination with AZD1222 (n = 18, left panel) or BNT162b2 (n = 18, right panel). All FACS analyses were on gated live monocytes. Asterisks indicate significant differences between time points. p-values resulting from one-way ANOVA and Tukey-Kramer multiple comparisons tests were < 0.0001 for AZD1222 and 0.0146 for BNT162b2 analyses. ***p < 0.001

Likewise, there were significant changes among granulocyte subpopulations and these were restricted to AZD1222-vaccinated subjects only (Supplementary Figs. 1 and 2). In detail, CD177^−^CD11b^+^ among CD14^+^CD16^−^ granulocytes were significantly elevated on days 2 and 13 following vaccination (Supplementary Fig. 1). By day 20, this subpopulation was still increased to some extent however, due to high variance, the difference to baseline did not reach statistical significance. Interestingly, dynamics of CD177^−^CD11b^−^ among CD14^+^CD16^+^ granulocytes followed an opposing trend with proportions being decreased on days 2 and 13 before returning to baseline by day 20 after vaccination (Supplementary Fig. 2). In summary, this expression data show that vaccination with BNT162b2 had almost no impact on the innate immune compartment, whereas vaccination with AZD1222 led to marked alterations in the compositions of granulocyte and monocyte subpopulations.

### 2.3 Changes among adaptive immune cell populations were most prominent after AZD1222-vaccination

In order to characterize the response of adaptive immune cells following vaccination, we next investigated the proportions of B- and T-lymphocyte subpopulations. Fig. 3A shows representative data of CD19^+^CD45RA^+^ B cells and illustrates for a participant vaccinated with AZD1222 a shift of subpopulations expressing CD27 and CD38, respectively. While CD27^+^CD38^bright^ plasmablasts were significantly enriched on day 6 following vaccination with AZD1222 and reached a median of 3.08%, a plasmablast peak after BNT162b2 vaccination was detectable on day 13, yet reached a median of 1.57% only (Fig. 3B, upper panels). The increase in plasmablasts after AZD1222 vaccination was flanked by an increase in CD27^+^CD38^−^ late memory B cells on days 13 and 20 (Fig. 3B, lower panels). No such alterations were observed after BNT162b2 vaccination.

**Fig. 3.**
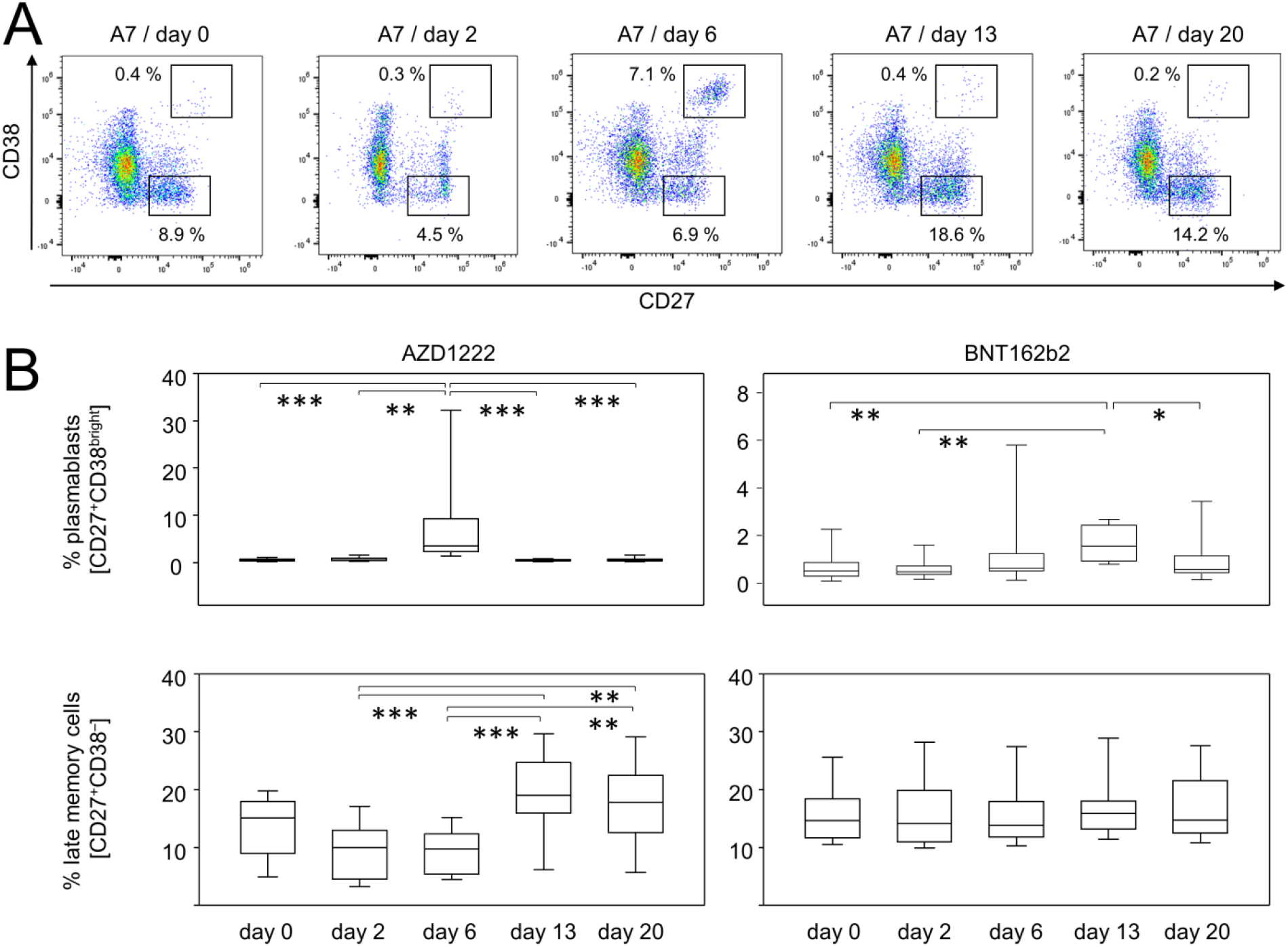
Vaccination with AZD1222 and BNT162b2 induced significant yet transient alterations among peripheral plasmablasts. AZD1222 also induced a significant increase in late memory B cells on days 13 and 20. **A** Pseudocolor plots for the expression of CD27 and CD38 on B cells are representative for the AZD1222 vaccination group. **B** Proportions of CD27^+^CD38^bright^ plasmablasts (top) and CD27^+^CD38^−^ late memory B cells (bottom) after vaccination with AZD1222 (n = 18, left) or BNT162b2 (n = 18, right). All FACS analyses were on CD19^+^CD45RA^+^ B cells. p-values resulting from one-way ANOVAs were < 0.0001 for both AZD1222 analyses. p-values resulting from Kruskal-Wallis and Dunn’s multiple comparisons tests were 0.0008 for the comparison of plasmablast and 0.6998 for the comparison of late memory B cells for the BNT162b2 analyses. *p < 0.05, **p < 0.01, ***p < 0.0001

Likewise, alterations among T cell subpopulations were restricted to AZD1222 vaccinees only (Supplementary Figs. 3 and 4). In detail, we detected a shift in the expression patterns of CD27 and CD38 on CD8^+^ cells especially two days after AZD1222 vaccination (Supplementary Fig. 3A). This translated into a significant enrichment of CD27^−^CD38^+^ terminally differentiated cytotoxic T cells for this group (Supplementary Fig. 3B). Supplementary Fig. 4A exemplifies for CD4^+^ cells a change in the expression patterns for CD27 and CD127 on day 20 after AZD1222 administration. We thus discovered that CD4^+^CD127^−^CD27^+^ effector memory T cells re-expressing RA were enriched towards the end of the observation period (Supplementary Fig. 4B). Taken together, we here demonstrated that significant changes among subpopulations of B- and T-lymphocytes were observed after vaccination with AZD1222 only.

### 2.4 AZD1222 and BNT164b2 vaccinations led to significantly different helper and cytotoxic T cell polarizations

So far we have shown that a single dose of AZD1222 was capable of significantly modifying immune cell compositions. However, we assumed that both vaccines would on a small scale induce a specific cellular immune response towards the SARS-CoV-2 spike protein, which would become detectable upon re-stimulation with the antigen. We therefore used both, recombinant spike protein and BNT162b2 vaccine and aimed to investigate cytokine profiles as well as the expression of inducible activation markers. In case of the recombinant spike protein, we expected it to be taken up by antigen presenting cells (APCs). Upon processing the protein in lysosomes, respective peptides would predominantly become displayed on human leukocyte antigen (HLA) class II molecules and thus, would be ready to activate spike protein specific T helper cells. By employing the mRNA vaccine, we anticipated its cellular uptake, translation into protein and then both, secretion for uptake by APCs and class II presentation as well as processing the protein for presentation via HLA class I molecules and thereby re-stimulating cytotoxic T cells^20^.

In order to establish a working protocol, we used PBMCs from fully vaccinated or COVID-19 convalescent blood donors and investigated the expression of the activation marker CD137 on unstimulated cells compared to cells challenged with either the recombinant spike protein or BNT162b2. Indeed, we found a significant activation of CD4^+^ but not CD8^+^ cells after providing the recombinant spike protein (Supplementary Fig. 5). In contrast, stimulation with the BNT162b2 vaccine led to a significant increase of CD137 expressing cells among both, CD4^+^ and CD8^+^ lymphocytes (Supplementary Fig. 5).

In a first approach, we used a classical ELISPOT assay in combination with recombinant spike protein to confirm for both vaccination regimes increasing amounts of IFNγ-secreting cells on day 20 compared to day 0. While Fig. 4A presents individual examples of ELISPOTs, Fig. 4B summarizes all results and indeed shows a significant increase in IFNγ-positive cells after BNT162b2 vaccination (p=0.0059). There was also a trend towards increased IFNγ-positive cells after AZD1222 vaccination however, this did not reach statistical significance (p=0.0968).

**Fig. 4.**
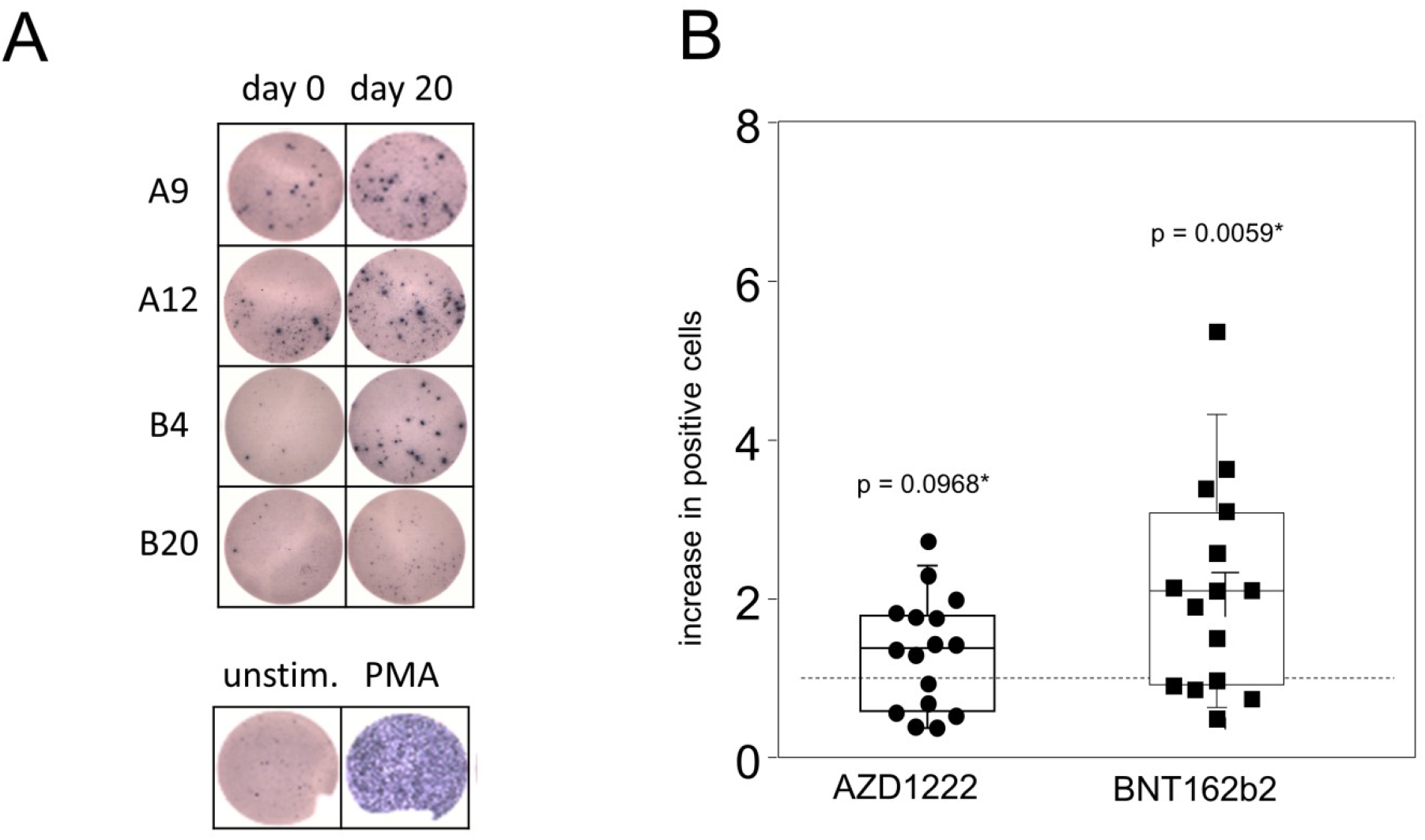
Spike specific IFNγ-producing lymphocytes were significantly increased after vaccination with BNT162b2. PBMCs were isolated at the day of vaccination (day 0) and 20 days later. Cells were then stimulated with recombinant SARS-CoV-2 spike protein for 24 h. Subsequently, an ELISPOT assay for the detection of IFNγ producing cells was performed. **A** Representative shots of spot forming cells from individuals vaccinated with either AZD1222 (A9 and A12) or BNT162b2 (B4 and B20). PBMCs were stimulated with PMA as a positive control (bottom). **B** The counts of spot forming cells from day 20 were normalized to counts from day 0. Data show the relative increase in IFNγ^+^ cells after vaccination (n = 16 for AZD1222 and n = 15 for BNT162b2). p-values result from one-sample t-tests assessing the differences of group means to the hypothetical value of 1.

We next sought to differentiate the AZD1222- and BNT162b2-induced adaptive cellular immune responses in more detail. Therefore, we cultured day 20 PBMCs in the presence or absence of BNT162b2 and surveyed activation and cytokine profiles by flow cytometry after Brefeldin A-capture of secretory proteins. Fig. 5A shows that following both vaccination regimes, re-challenge with spike mRNA led to significantly increased amounts of CD8^+^CD137^+^ T cell that also expressed CD25 (IL-2Rα), suggesting the differentiation to an effector phenotype. In addition, both vaccines facilitated the expansion of inducible spike specific cytotoxic effector T cells as demonstrated by significantly increased percentages of FasL^+^CD8^+^ cells after *in vitro* re-stimulation (Fig. 5B). We further detected a significant increase in IFNγ-producing CD8^+^ T cells from AZD1222-but not from BNT162b2-vaccinated donors (p = 0.0325 vs 0.1514, Fig. 5C). Finally, re-stimulation with spike mRNA decreased IL-2^+^IL-10^+^ co-expressing regulatory CD8^+^ cells for both AZD1222 and BNT162b2, the latter short of reaching statistical significance (p = 0.0058 vs 0.0768, Fig. 5D).

**Fig. 5.**
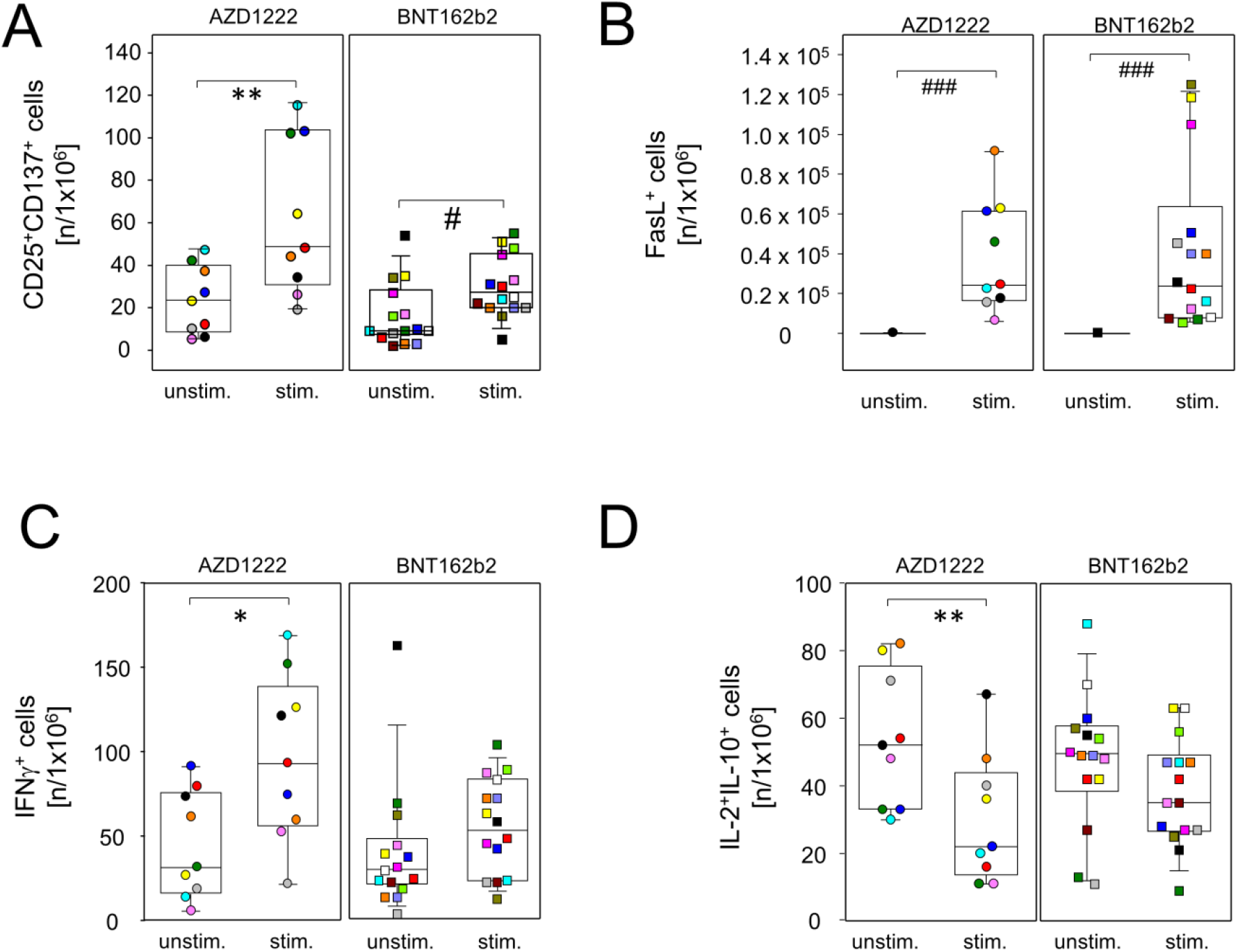
Vaccination with single doses of AZD1222 or BNT162b2 induced the expansion of cytotoxic effector T cells. PBMCs were isolated on day 20 after vaccination and stimulated with (stim.) or without (unstim.) spike protein encoding mRNA (BNT162b2) for 24 h and analyzed by flow cytometry for the expression of inducible activation markers and intracellularly trapped cytokines among CD8^+^ cells. Sample sizes where n = 9 for AZD1222 and n = 15 for BNT162b2, respectively. Paired samples are illustrated by color coding. **A**,**B** *in vitro* re-stimulation significantly increased CD25^+^CD137^+^ (**A**) and FasL expressing cells in both vaccination groups (**B**). **C**,**D** INFγ– producers were increased (**C**) and IL-2 and IL-10 co-expressing cells were significantly reduced among AZD1222 vaccinees only (**D**). *p < 0.05, **p < 0.01 resulting from paired t-tests. ^#^p < 0.05, ^###^p < 0.001 resulting from Wilcoxon signed rank tests for matched pairs.

Significantly increased amounts of T helper cells with an effector phenotype (CD4^+^CD25^+^CD137^+^) were also detected for both vaccine regimens after *in vitro* re-stimulation (Fig. 6A). In contrast, an increase in pro-inflammatory helper T cells producing TNFα was restricted to re-stimulated cultures from day 20 AZD1222 donors (Fig. 6B). Of note, neither vaccination induced the expansion of spike specific type 2 helper T cells as demonstrated by a lack of inducible IL-4 production by CD4^+^ cells after re-stimulation (Fig. 6C). Similar to regulatory CD8^+^ cells and again restricted to the AZD1222 group, re-stimulation with spike mRNA statistically significantly reduced anti-inflammatory IL-2 and IL-10 co-production in a subset of CD4^+^ cells (Fig. 6D).

**Fig. 6.**
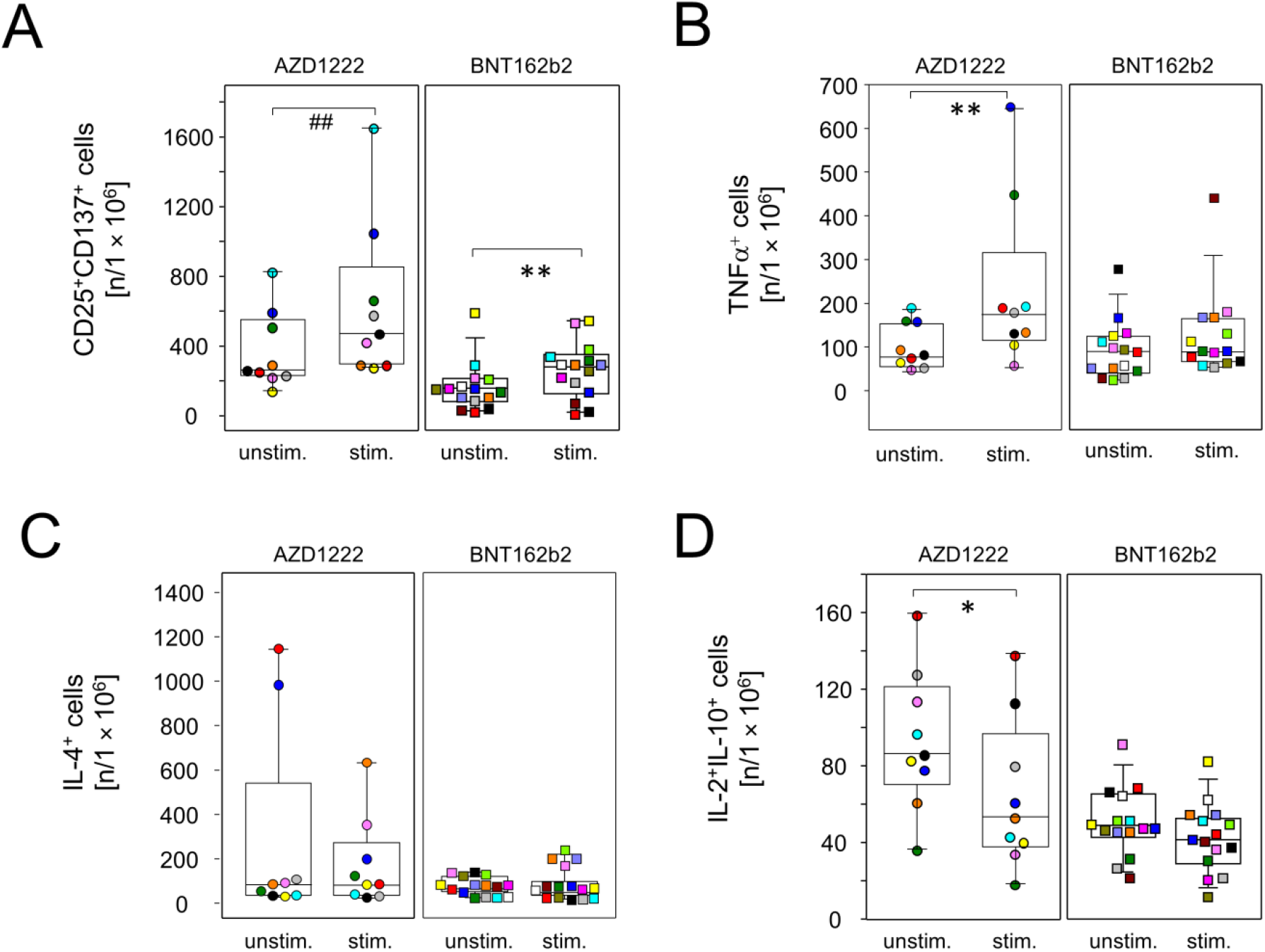
AZD1222 vaccination supported the induction of type 1 helper T cells. PBMCs were isolated on day 20 after vaccination and stimulated with (stim.) or without (unstim.) spike protein encoding mRNA (BNT162b2) for 24 h and analyzed by flow cytometry for the expression of inducible activation markers and intracellularly trapped cytokines among CD4^+^ cells. Sample sizes where n = 9 for AZD1222 and n = 15 for BNT162b2, respectively. Paired samples are illustrated by color coding. **A**,**B** *in vitro* re-stimulation significantly increased CD25^+^CD137^+^ cells in both vaccination groups (**A**) and TNFα-producing cells in the AZD1222 vaccinated group, only (**B**). **C** Neither vaccination allowed for the re-stimulation of IL-4-producing cells. **D** IL-2 and IL-10 co-expressing cells were significantly reduced among AZD1222 vaccinees only. *p < 0.05, **p < 0.01, resulting from paired t-test. ^##^p < 0.01, resulting from Wilcoxon signed rank test for matched pairs.

In summary, these results demonstrate that single doses of either AZD1222 or BNT162b2 induced the polarization of spike specific CD4^+^ and CD8^+^ effector T cells. More pronounced changes were again observed after vaccination with AZD1222 that included reduced proportions of IL-2 and IL-10 co-expressing CD4^+^ and CD8^+^ cells alike, combined with increased IFNγ-producing CD8^+^ and TNFα-producing CD4^+^ cells.

### 2.5 BNT162b2 vaccination led to significantly more spike protein-specific plasma IgG and IgA

Even though both vaccines led to significant adaptive immune activation, alterations to inducible effector functions followed distinct patterns for AZD1222 and BNT162b2, respectively. We therefore sought to investigate whether these differences resulted in the production of diverse collections of immunoglobulin isotypes. To that extent, spike protein specific IgM, IgG and IgA were assessed for all time points via ELISA. As shown in Fig. 7, either vaccine induced the production of detectable amounts of antibodies as early as day 13. However, significant differences emerged between AZD1222 and BNT162b2 vaccination concerning the distribution and amounts of spike specific antibody isotypes. In detail, AZD1222 predominantly induced IgM and IgG, while IgA was virtually absent, even at day 20 after vaccination. In contrast, although not significantly different from AZD1222, BNT162b2 elicited little IgM. There were though increased IgG and IgA titers in BNT162b2 vaccinated subjects and these differences reached significances on day 20 for IgG and already on day 13 for IgA (Fig. 7). When looking at response rates instead of Ig titers, there were significantly more IgM and significantly less IgA responders to AZD1222 compared to BNT162b2 as calculated via Fisher’s exact test (Supplementary Table 3). Collectively, disparate vaccine strategies for spike protein delivery impacted differently on the humoral immune response and shaped distinctive antibody isotype layouts after single doses of AZD1222 and BNT162b2.

**Fig. 7.**
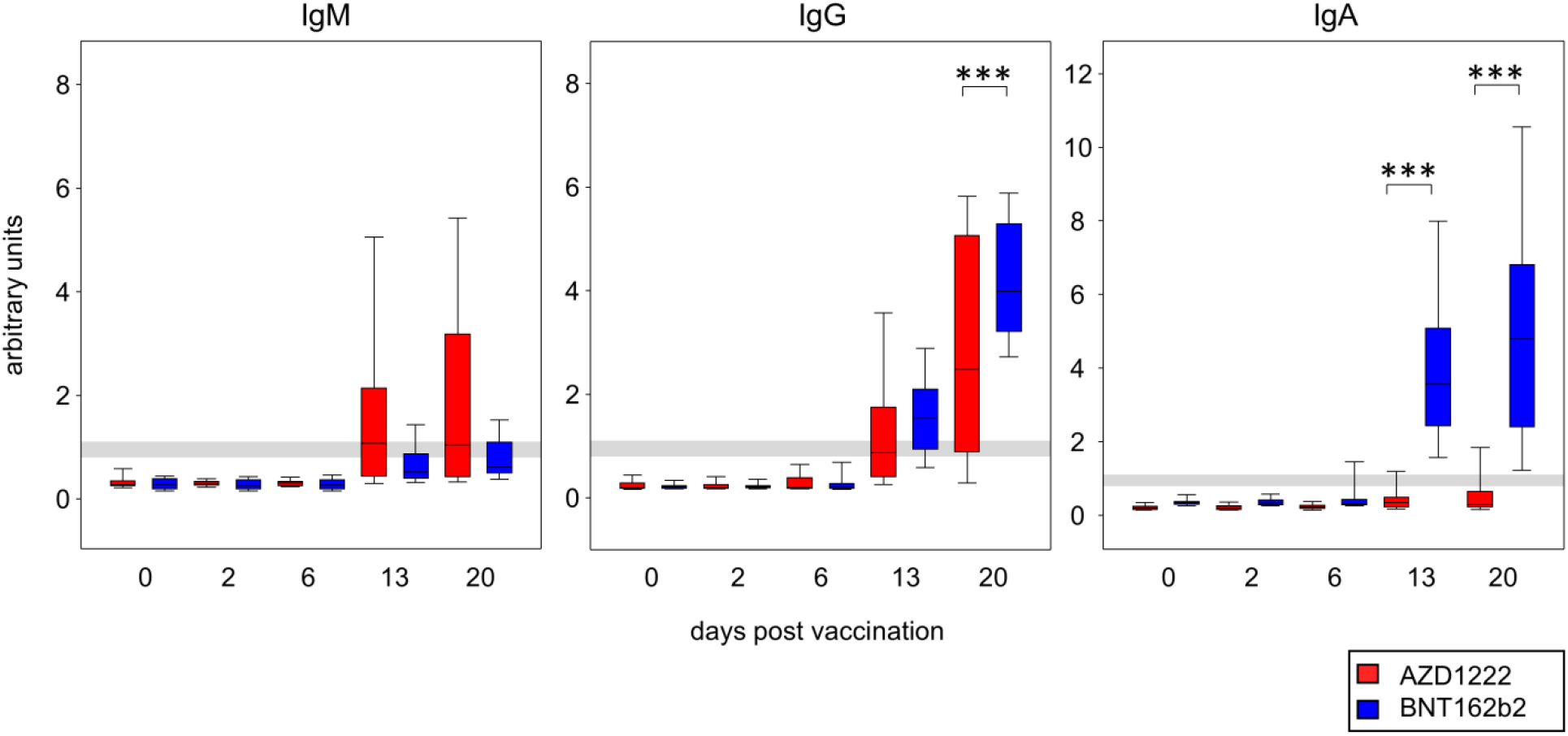
A single dose of BNT162b2 induced significantly higher amounts of spike protein binding IgG and IgA than AZD1222. Antibodies were detected by ELISA and absorbance readouts were normalized to calibrator values to obtain arbitrary units. Grey lines depict the positive response threshold within a range of 0.8 to 1.1 arbitrary units. Sample sizes were n = 18 for AZD1222 and n = 18 for BNT162b2, respectively. p-values resulting from one-way ANOVA and Tukey-Kramer multiple comparisons tests were < 0.0001 for all three isotype analyses. ***p < 0.001

## 3 Discussion

We here compare early immune reactions to the primary vaccination against SARS-CoV-2 with either AZD1222 from AstraZeneca or BNT162b2 from BioNtech^6,8^. While both vaccines elicit strong cellular and humoral responses, the individual impacts on the peripheral immune compartment were strikingly different. In detail, the adenoviral vector AZD1222 led to a transient yet profound leukopenia on day 2, involving significant reductions of B and T lymphocytes as well as granulocytes. As decreased leukocyte counts have previously been reported for both, regular adenoviral infections^21^ and adenovirus-mediated gene therapeutic approaches^22,23^, we consider it likely that the leukopenia observed here can be attributed to the viral vector rather than the spike protein^24^.

Likewise, the AZD1222-induced changes among peripheral immune cell proportions are reminiscent of viral infections. For instance, short-term enrichments of pro-inflammatory monocytes have been observed in patients infected with Dengue or human immunodeficiency virus, respectively^25,26^. Both, the enrichment of CD4^+^ effector memory T cells re-expressing RA (TEMRA) and terminally differentiated cytotoxic CD8^+^ T cells have been associated with human cytomegalovirus^27–29^. In addition, the class switched, late memory B cells that we observed after AZD1222 vaccination have previously been associated with an efficient control of viral infections^30,31^. Together, our data suggest that it is the adenoviral vector rather than the encoded spike protein that elicits a significant immune cell response in the periphery.

By comparison, the immune cell response to BNT162b2 vaccination appeared rather bland and thus corroborated the observation that this vaccine is globally well tolerated among first dose recipients^9^. Indeed, the BNT162b2 encoded mRNA bears an m1Ψ modification which attenuates innate immune sensing^32^. We therefore assume that, the relatively small enrichment of peripheral plasmablasts after BNT162b2 vaccination resulted from an adaptive response towards the SARS-CoV-2 spike protein whereas the larger enrichment observed with AZD1222 likely reflects a combined response towards the spike protein and the adenoviral vector.

When comparing the specific antibody responses against the spike protein, we did not observe any quantitative differences in total Ig between both vaccines. However, the isotypes produced were significantly different with AZD1222 inducing primarily IgM and IgG compared to predominantly IgG and IgA by BNT162b2. Even though we cannot yet predict whether this trend will be continued beyond the first three weeks after vaccination, a pronounced IgA response following BNT162b2 vaccination may explain its superior effectiveness in preventing symptomatic COVID-19 after both, infection with wild type SARS-CoV-2 and its variants^15,17,19,33^. Optimizing existing vaccines might therefore aim at alternative antigen delivery, e.g. towards mucosal sites, in order to not only support IgA production but also tissue-resident effector cells which will help contain viral loads at the nasopharyngeal entry sites^34,35^.

Optimizing vaccines may also aim at modifying potential bystander effects. When analyzing the spike protein-specific T cell responses, we observed that both vaccines elicited functional immune responses. Both vaccination regimes expanded effector cells to a comparable degree as documented by significant increases in activated CD25 and CD137 co-expressing CD4^+^ and CD8^+^ T lymphocytes as well as FasL expressing CD8^+^ cells upon re-challenge. However, when looking at intracellular cytokine production, AZD1222 induced a prominent Th1 response as illustrated by significant increases in IFNγ and TNFα, respectively. Because adenoviral vectors have previously been shown to facilitate strong cellular immunity towards the delivered antigen and drive the expansion of type 1 helper (Th1) cells^6^, we believe that this Th1 reaction towards the adenoviral vector exerted some bystander effect on the response against the spike protein. However, an inordinate Th1 response may foster a cytokine layout that is hardly supportive of class-switch recombination towards IgA^36^. On the other hand, we assessed neither IL-5 nor TGFβ and are therefore not yet able to verify whether BNT162b2 indeed induced more IgA promoting cytokines.

Of note, our ELISPOT experiments addressing spike protein-specific Th1 cells before and after vaccination found an even stronger induction of IFNγ-producing T helper cells among BNT162b2 compared to AZD1222 vaccinees. Even though these results seemingly contradict the intracellular cytokine readout of stimulated vs unstimulated day 20 T cells, they show that both vaccines induced Th1 responses. In addition, we found a reduction of IL-2 and IL-10 co-producing CD4^+^ and CD8^+^ T cells upon *in vitro* re-stimulation which however, only reached significance in the case of AZD1222 vaccination. IL-10 is a hallmark of regulatory T cells and exerts anti-inflammatory effects via suppressing not only effector T cells, but also antigen presentation and the secretion of inflammatory cytokines by APCs^37–39^. We like to speculate that this significant reduction of IL-10 expression is not restricted to *in vitro* re-challenge but may also occur after booster immunization and perpetuate a Th1 response that impedes an IgA promoting cytokine milieu.

This study has a few limitations, among them the small samples sizes. Nonetheless, our results depict for both vaccines significantly disparate effects on the peripheral immune layout and on the regulation of T cell effector molecules. Another limitation is the lack of assaying neutralizing SARS-CoV-2 specific antibodies. However, previous studies have already demonstrated that total spike-binding Ig titers strongly correlate with the amounts of neutralizing antibodies and are therefore a suitable measure for humoral protection from SARS-CoV-2 infection^5,40–42^. In summary, we consider the description of disparate vaccine effects on the immediate immune response the strength of our study and we believe that our results will be of use for further optimization of vaccination strategies.

## 4 Materials & Methods

### 4.1 Study participants and blood sampling

Study participants were recruited from the coordination center for clinical studies at the Rostock University Medical Center. Individuals with a study-independent appointment at a vaccination center for vaccination with either AZD1222 or BNT162b2 were eligible to participate. Blood samples were obtained by consecutive venipuncture on the day of vaccination (d0) and on days two, six, thirteen and twenty thereafter. Peripheral blood mononuclear cells (PBMCs) were isolated from anti-coagulated blood by density gradient centrifugation using Ficoll-Paque™ PLUS to the manufacturer’s instructions (Cytiva, Marlborough/MA, United States). PBMCs were subsequently suspended in fetal calf serum (FCS, Thermo Fisher, Waltham/MA, United States) containing 10% dimethyl sulfoxide (Sigma-Aldrich, St. Louis/MO, United States) and were frozen at -70 °C until further use. Plasma samples were obtained by centrifugation of anti-coagulated blood and were frozen afterwards. This study was approved by the ethics committee of the Rostock University Medical Center under the file number A 2020-0086. Written informed consent was provided by all participants.

### 4.2 Flow cytometric analyses of surface markers

For the analysis of surface expression markers, 100 µL of anticoagulated whole blood were used. In order to reduce unspecific antibody-conjugate binding, 10 µL FCS, 5 µL True-Stain Monocyte Blocker™ and 5 µL anti-Fc receptor TruStain FcX™ (Biolegend, San Diego/CA, United States) were added and incubated for 15 min on ice. The following amounts of antibody:fluorophore-combinations were used: 0.25 µg CD127:APC/R700 (clone HIL-7R-M21), 1 µg CD147:BV421 (TRA-1-85), 0.5 µg CD45RO:BV480 (UCHL1, BD Biosciences, Franklin Lakes/NJ, United States), 1 µg CD11b:PerCP/Cy5.5 (ICRF44), 0.8 µg CD11c:BV785 (3.9), 0.56 µg CD14:BV510 (63D3), 0.13 µg CD16:BV650 (3G8), 0.06 µg CD19:APC/Fire810 (HIB19), 0.13 µg CD20:SparkNIR685 (2H7), 0.5 µg CD27:BV605 (O323), 0.25 µg CD3:SparkBlue550 (SK7), 0.25 µg CD304:AlexFluor647 (12C2), 0.03 µg CD4:BV750 (SK3), 0.5 µg CD45RA:APC/Fire750 (HI100), 0.13 µg CD56:BV711 (5.1.H11), 0.13 µg CD8:BV570 (RPA-T8), 0.5 µg CD95:PE/Cy5 (DX2), 0.13 IgD:PE/Dazzle594 (IA6-2), 0.13 µg PD-1:APC (A17188B, Biolegend), 0.06 µg CD38:PerCP/eFluor710 (HB7, Thermo Fisher), 0.06 µg CD177:PE/Vio770 (REA258), 0.05 µg CD25:PE (REA570, Miltenyi, Bergisch Gladbach, Germany). Antibodies were incubated for 15 min on ice in the dark. Subsequently, Apotracker™ Green (Biolegend) was added according to the manufacturer’s instruction without washing, followed by incubation for 30 min on ice. In order to lyse erythrocytes, 2.2 mL Fixative-Free Lysing Solution (Thermo Fisher) was added and incubated for 20 min at room temperature. Subsequently, 0.03 µg 4’,6-diamidino-2-phenylindole (DAPI, Biolegend) were added as a live/dead discriminator and incubated for 5 min. Finally, data acquisition was performed on the Cytek^®^ Aurora flow cytometer running on the SpectroFlo Software version 2.2.0.3 (Cytek Biosciences, Fremont/CA, United States). Analysis of flow cytometry data was done using FlowJo software version 10.7 (FlowJo, Ashland/OR, United States). The gating scheme is shown in Supplementary Fig. 6. Dimension reduction of down-sampled (5 × 10^4^ live cells per sample) and concatenated data sets was performed using the FlowJo plugin for the algorithm “uniform manifold approximation and projection” (UMAP)^43^.

### 4.3 Interferon gamma ELISPOT

PBMCs from d0 and d20 were thawed, centrifuged and suspended in complete RPMI medium containing 10% FCS, 1% Penicillin/Streptomycin, 2 mM L-Glutamine (Thermo Fisher), 10 mM HEPES and 1 mM sodium pyruvate (PAN-Biotech, Aidenbach, Germany). Cell counts were determined cytometrically on the Cytek^®^ Aurora (Cytek Biosciences) using DAPI (Biolegend) as a live/dead discriminator. Five hundred thousand PBMCs were pipetted into a 96-well U-bottom plate and centrifuged for 5 min at 4 °C and 400 × *g*. Subsequently, supernatants were removed by carefully blotting the plate on a paper tissue. Cells were then suspended in 36 µL complete RPMI medium containing 0.2 µg of the SARS-CoV-2 trimeric spike protein (R&D Systems, Minneapolis/MN, United States). Afterwards, PBMCs were transferred into a 96-well enzyme-linked immune absorbent spot (ELISpot) assay plate coated with capture antibodies specific for human interferon (IFN)γ (R&D Systems). After incubating the cells for 30 min at 37 °C, 164 µL of complete RPMI medium was added to all wells followed by 24 h incubation at 37 °C in a CO_2_ incubator (Binder, Tuttlingen, Germany). The ELISpot assay was then performed according to the manufacturer’s guidelines. The numbers of IFNγ producing cells were determined by automated counting using the ImmunoSpot^®^ analyzer running on the ImmunoSpot^®^ Software version 5.0.9.15 (CTL Europe, Bonn, Germany). The counts of IFNγ-positive cells were normalized to their respective paired sample from d0.

### 4.4 T cell re-stimulation and intracellular cytokine staining assay

For the establishment of T cell re-stimulation and intracellular cytokine staining assays, blood was collected from six fully vaccinated subjects and one patient who had recovered from COVID-19 at least 2 weeks after the second vaccination and infection, respectively. PBMCs were isolated and frozen until further use in the same fashion as described above. For assaying the vaccinated study participants, day 20 PBMCs were used. Upon thawing, PBMCs were counted as described above and aliquots of 0.8 million were seeded into single wells of 96-well U-bottom plates. Every sample was stimulated at least in duplicates. After centrifugation, cells were re-suspended and stimulated in a total volume of 36 µL complete RPMI medium with either 1 µg of the BNT162b2 vaccine (BioNTech, Mainz, Germany) or with 0.2 µg of the SARS-CoV-2 trimeric spike protein or left without any stimulation. PMA (10 ng/ mL) and Ionomycin (1 µg/mL) stimulated samples were processed in parallel as positive controls. After adding 164 µL of complete RPMI medium, cells were incubated for 20 h under 5 % CO_2_ atmosphere at 37°C. One microgram of Brefeldin A (Sigma-Aldrich) was added thereafter followed by incubation for another 4 h. Successive incubation steps were performed in the dark. Duplicate samples were pooled, washed in PBS (Thermo Fisher), suspended in PBS containing 2,000-fold diluted ZombieNIR dye (Biolegend) and incubated for 20 min at room temperature. Thereafter, cells were washed and suspended in autoMACS^®^ Running Buffer (RB, Miltenyi Biotec, Bergisch Gladbach, Germany). Subsequently, unspecific antibody-conjugate binding sites were blocked by adding FCS, True-Stain Monocyte Blocker™ and anti-Fc receptor TruStain FcX™ (Biolegend) for 10 min at room temperature. Surface antigens were stained by incubating the cells with following antibody:fluorophore-combinations: 1.25 µg CD3:FITC (clone UCHT1), 0.02 µg CD4:BV750 (SK3), 0.06 µg CD8:BV570 (RPA-T8), 0.5 µg Fas-L:PE (NOK1), 1 µg CD25:APC (BC96, Biolegend), 1.25 µL CD127:APC/R700 (HIL-7R-M21) and 0.25 µg CD137:BV480 (4B4-1, BD Biosciences) for 15 min at room temperature. Cells were subsequently washed in RB, suspended in 100 µL Fixation Buffer (Biolegend) and incubated at room temperature for 20 min. Cells were washed twice and then suspended in Intracellular Staining Permeabilization Wash Buffer (Biolegend). After blocking unspecific binding sites as described above, 0.5 µg Granzym B:AlexaFluor647 (clone 6B11), 2.5 µg IFNγ:PerCP/Cy5.5 (4S.B3), 0.63 µg IL-2:BV650 (MQ1-17H12), 0.3 µg IL-4:PE/Dazzle594 (MP4-25D2), 1 µg IL-10:BV421 (JES3-907, Biolegend) and 0.13 µg TNFα:PE/Cy7 (Mab11, Thermo Fisher) were added and incubated for 30 min at room temperature. Finally, cells were washed twice in RB. Data acquisition and analysis of expression data was performed as described above. The gating scheme for the intracellular cytokine staining assay is shown in Supplementary Fig. 7.

### 4.5 SARS-CoV-2 Trimeric Spike Specific Antibodies

Plasma samples from all time points were thawed on ice and centrifuged at 10,000 × *g* in order to remove precipitates. For the detection of SARS-CoV-2 trimeric spike protein specific immunoglobulin (Ig)G and IgA levels, plasma was diluted 101-fold. The enzyme-linked immunosorbent assays (ELISA) for these isotypes was conducted after the manufacturer’s specifications (Euroimmun, Lübeck, Germany). In order to determine IgM levels, plasma samples were diluted 1,000-fold and the ELISA was performed to the manufacturer’s instructions (Thermo Fisher). The absorbance was detected at 450 nm (A450) on the Infinite^®^ 200 automated plate reader (Tecan, Männedorf, Switzerland). Absorbance readouts were normalized to calibrator values and were reported as arbitrary units. According to the manufacturer’s guidelines, calibrated sample values between 0.8 and 1.1 were considered borderline and above 1.1 were considered clearly positive. Samples from individuals with arbitrary unit values of less than 0.8 were considered non-responders.

### 4.6 Statistical Analysis

Data analyses were performed using R (version 3.5.1) and InStat version 3.10 (GraphPad, San Diego/CA, United States). Data sets were evaluated for Gaussian distribution using the Kolmogorov-Smirnov test. Under the assumption of normally distributed sample data, multiple independent groups were compared by one-way analysis of variance (ANOVA) followed by post-hoc pairwise comparisons using the Tukey-Kramer test. Data, which did not following Gaussian distribution, was compared by the Kruskal-Wallis one-way analysis of variance combined with Dunn’s test for multiple comparisons. The one-sample t-test was performed to compare single group means to a hypothetical value. Differences between dependent samples were assessed by the paired t-test given normal distribution and by the Wilcoxon signed rank test in case of deviation from Gaussian distribution. A p-value of less than 0.05 was considered statistically significant. Data visualization was performed with SigmaPlot version 13.0 (Systat Software GmbH, Erkrath, Germany).

## Data Availability

All data generated or analyzed during this study are included in this published article (and its supplementary information files).

## Acknowledgements

We thank all our participants for taking part in this study and providing blood samples. The authors want to particularly thank Wendy Bergmann (Core Facility for Cell Sorting and Cell Analysis, Rostock University Medical Center) for her help during flow cytometry panel design and setup. Organizational and documentary support was provided by the coordination center for clinical studies and Manja Ehmke (Division of Tropical Medicine and Infectious Diseases, Center of Internal Medicine II) from Rostock University Medical Center. Blood samples were also collected by Anxhela Muca and Stefanie Brigitte Amann-Stegbauer (both Core Facility for Cell Sorting and Cell Analysis, Rostock University Medical Center). This study was financially supported by the Federal State of Mecklenburg-Western Pomerania via the “Sondervermögen des MV Schutzfonds Säule Gesundheit”.

## Author contributions

M.M., B.M-H. and E.C.R. designed the experiments. B.M-H., B.S., S.M. and M.S. recruited vaccinees and documented their data. B.S. and S.M. collected peripheral blood samples. M.M and J.V performed flow cytometry, immunsorbent and immunospot assays, executed intracellular cytokine staining assays, processed peripheral mononuclear cells and analyzed the data. B.M-H. and J.V. performed statistical analyses. B.M-H. and J.V. wrote the first draft of the manuscript. M.M. wrote sections of the manuscript. All authors contributed in manuscript revision, read and approved the submitted version.

