## Supplementary Information for "Single-Dose SARS-CoV-2 Vaccination With BNT162b2 and AZD1222 Induce Disparate Th1 Responses and IgA Production"

Michael Müller, Johann Volzke, Behnam Subin, Silke Müller, Martina Sombetzki, Emil C. Reisinger, Brigitte Müller-Hilke

**Contents**

Supplementary Figures 1-7

Supplementary Tables 1-3

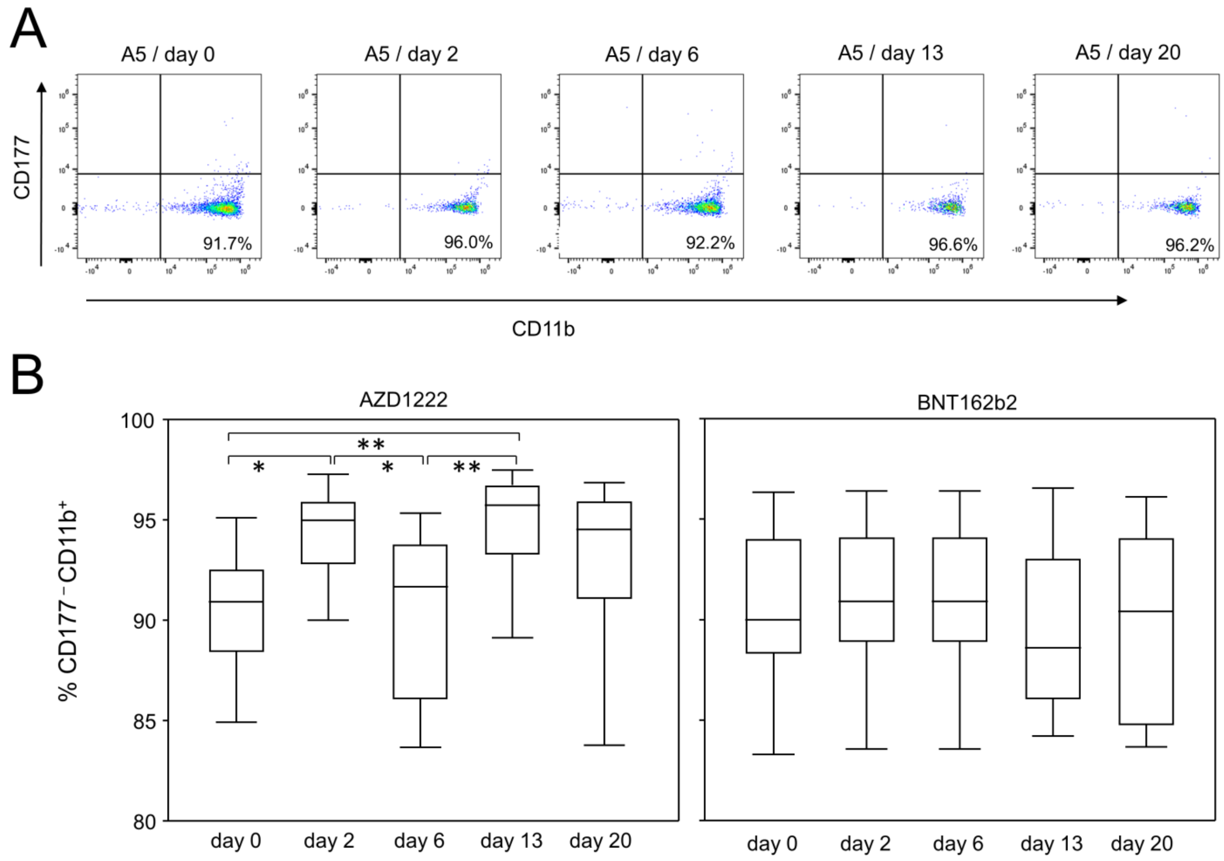


**Supplementary Fig. 1 Vaccination with AZD1222, unlike BNT162b2, led to an intermittent increase in $CD14^{+}CD16^{-}CD11b^{+}CD177^{-}$ granulocytes. A** Pseudocolor plots for the expression of CD11b and CD177 on $SSC^{hi}CD14^{+}CD16^{-}$ granulocytes are representative for the AZD1222 vaccination group. **B** Proportions of $CD177^{-}CD11b^{+}$ granulocytes after vaccination with AZD1222 (n = 18, left panel) or BNT162b2 (n = 18, right panel). p-values resulting from Kruskal-Wallis and Dunn´s multiple comparisons tests were 0.0004 for AZD1222 and 0.6760 for BNT162b2 analyses, respectively. *p < 0.05, **p < 0.01

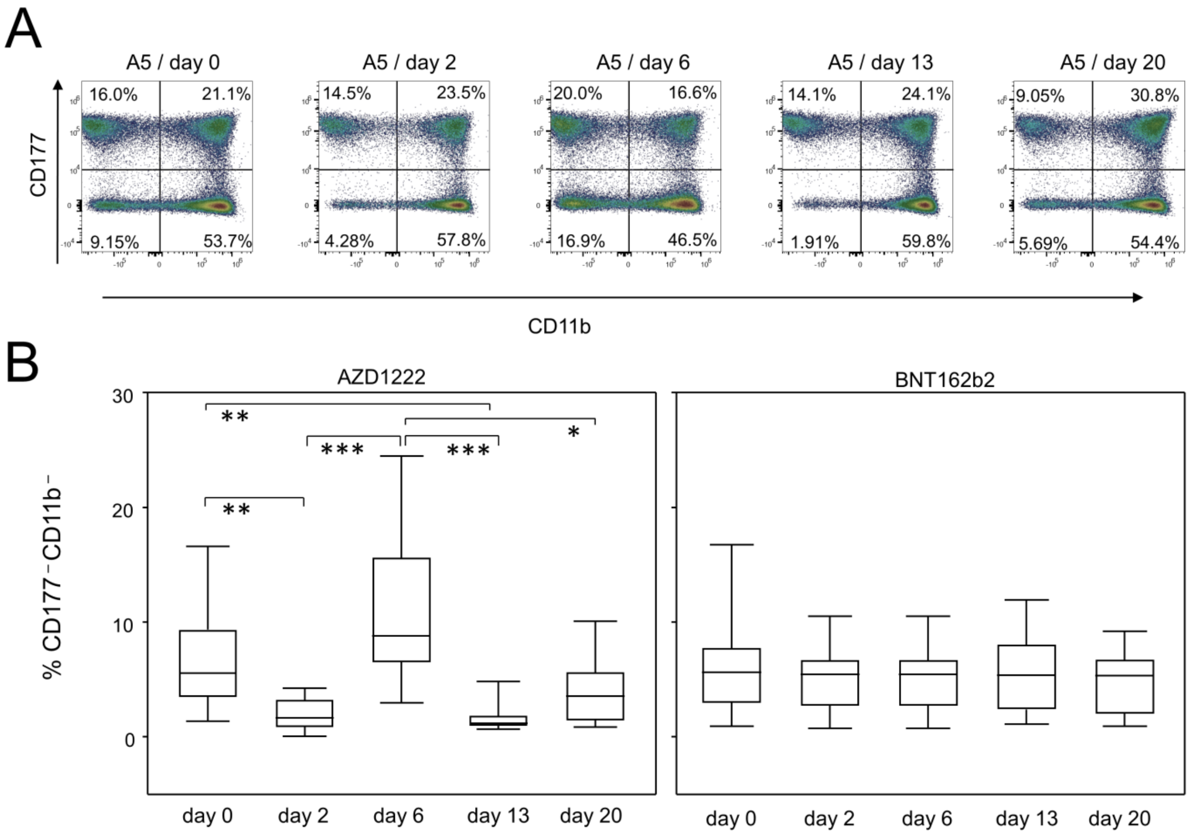


**Supplementary Fig. 2 $CD14^+CD16^+CD11b^-CD177^-$ granulocytes were decreased after vaccination with AZD1222 only. A** Pseudocolor plots for the expression of CD11b and CD177 on $SSC^{hi}CD14^+CD16^+$ granulocytes are representative for the AZD1222 vaccination group. **B** Proportions of $CD177^-CD11b^-$ granulocytes after vaccination with AZD1222 (n = 18, left panel) or BNT162b2 (n = 18, right panel). FACS analyses were gated on $CD14^+CD16^+$ granulocytes. p-values resulting from Kruskal-Wallis and Dunn´s multiple comparisons tests were < 0.0001 for AZD1222 and 0.9152 for BNT162b2 analyses, respectively. *p < 0.05, **p < 0.01, ***p < 0.001

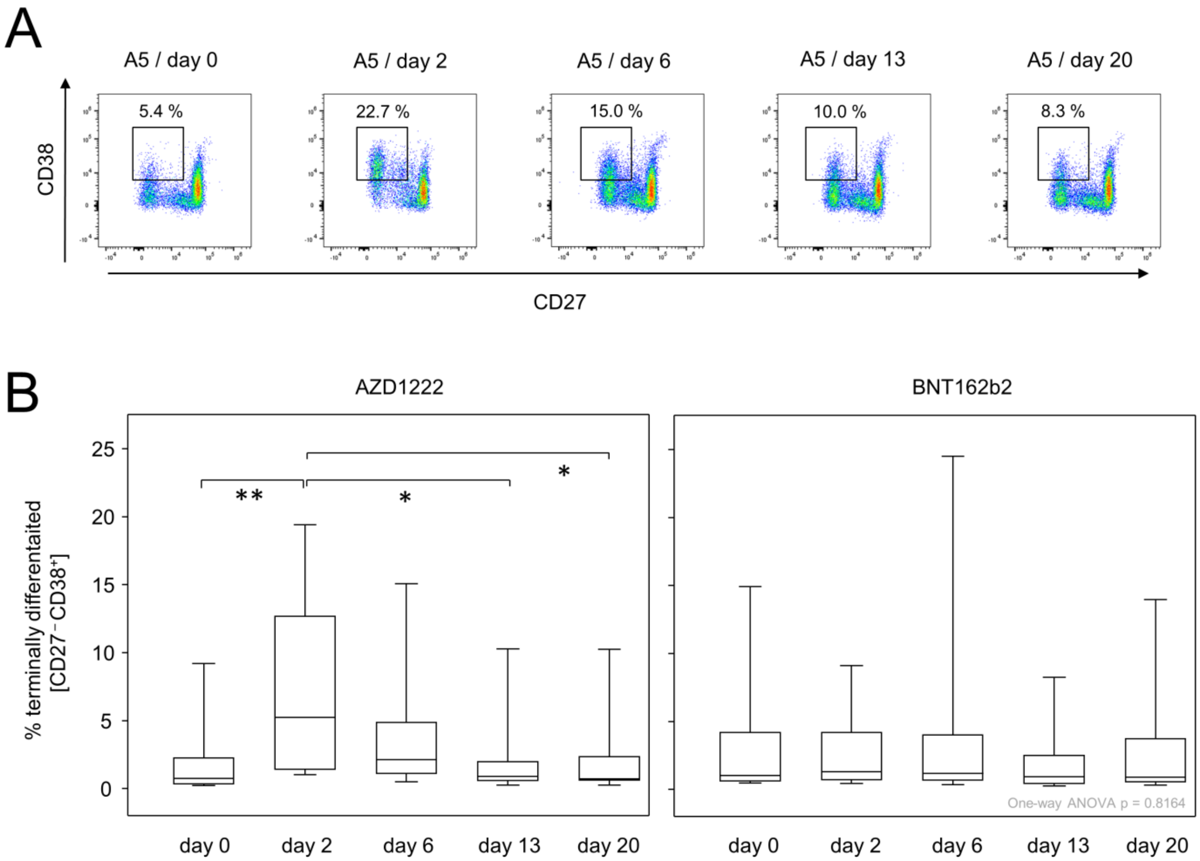


**Supplementary Fig. 3 AZD1222 vaccination induced the transient increase of $CD27^{-}CD38^{+}$ terminally differentiated $CD8^{+}$ T cells. A** Pseudocolor plots for the expression of CD27 and CD38 on $CD8^{+}$ T cells are representative for the AZD1222 vaccination group. **B** Proportions of $CD8^{+}CD27^{-}CD38^{+}$ T cells after vaccination with AZD1222 (n = 18, left panel) or BNT162b2 (n = 18, right panel). All FACS analyses were on $CD8^{+}$ T cells. p-values resulting from Kruskal-Wallis and Dunn´s multiple comparisons tests were 0.0009 for AZD1222 and 0.7905 for BNT162b2 analyses, respectively. *p < 0.05, **p < 0.01

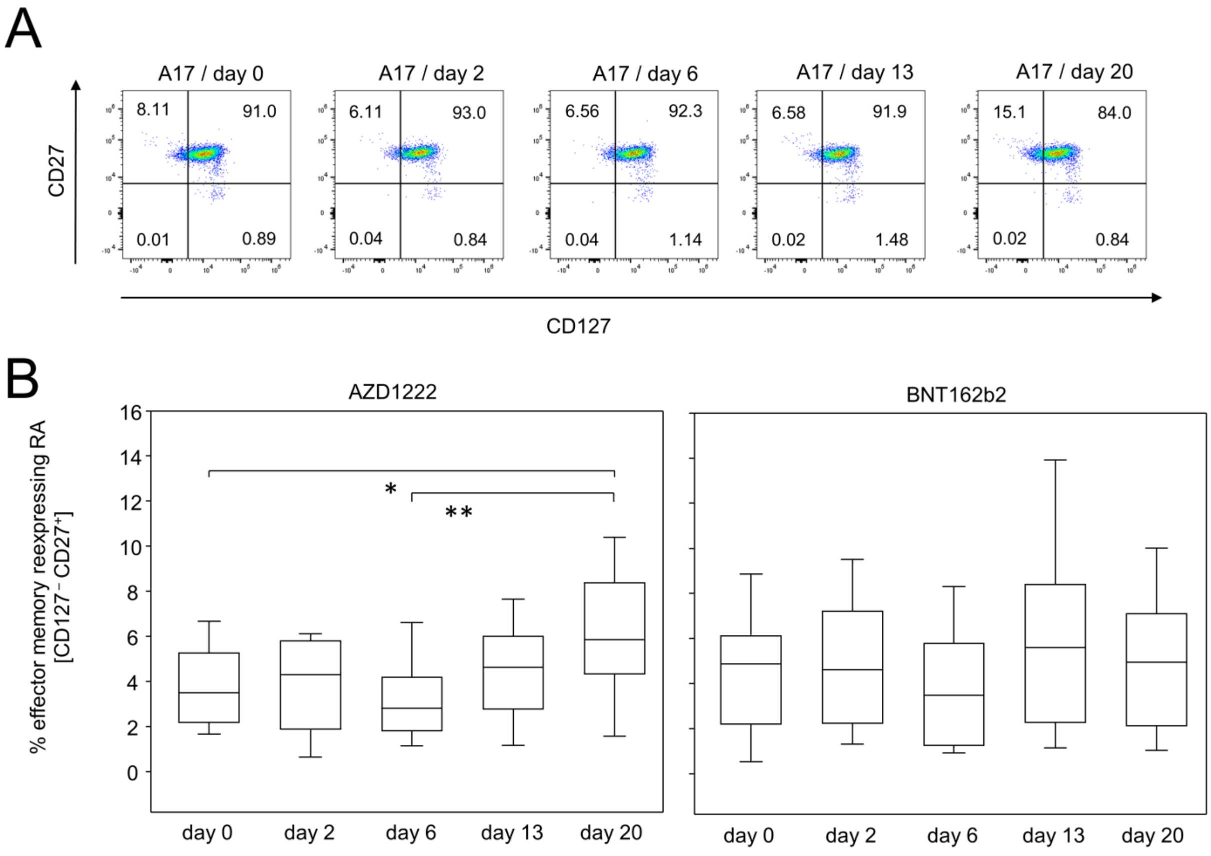


**Supplementary Fig. 4 CD4$^+$ effector memory T cells re-expressing RA were increased towards the end of the observation period after AZD1222 vaccination. A** Pseudocolor plots for the expression of CD127 and CD27 on CD4$^+$ T cells are representative for the AZD1222 vaccination group. **B** Proportions of CD4$^+$CD127$^+$CD27$^+$ T cells after vaccination with AZD1222 (n = 18, left panel) or BNT162b2 (n = 18, right panel). All FACS analyses were on CD4$^+$ T cells. p-values resulting from one-way ANOVA and Tukey-Kramer multiple comparisons tests were 0.0063 for AZD1222 and 0.4955 for BNT162b2 analyses, respectively. *$p < 0.05$, **$p < 0.01$

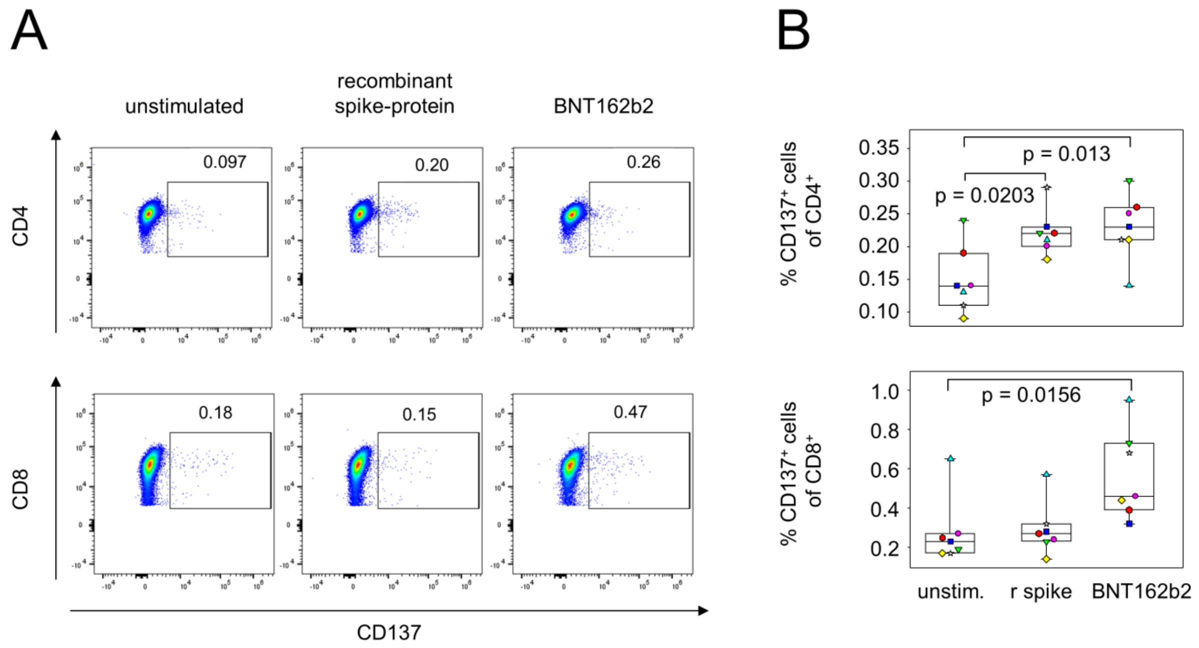


**Supplementary Fig. 5 Establishment of a protocol for the re-stimulation of SARS-CoV-2 spike protein specific immune cells.** PBMCs were isolated from fully vaccinated (n = 6) and COVID-19 convalescent (n = 1) blood donors and were in vitro re-stimulated with either recombinant spike protein (r spike) or the spike protein encoding mRNA (BNT162b2). **A** Representative pseudocolor plots showing the expression of CD137 on $CD4^+$ (upper panel) or $CD8^+$ T cells (lower panel) exemplifies activation. **B** Quantitative data for different stimulation regimen show that recombinant spike protein induced the enrichment of activated $CD4^+$ T cells, whereas spike protein encoding mRNA was able to increase the proportions of both activated $CD4^+$ and $CD8^+$ T cells.

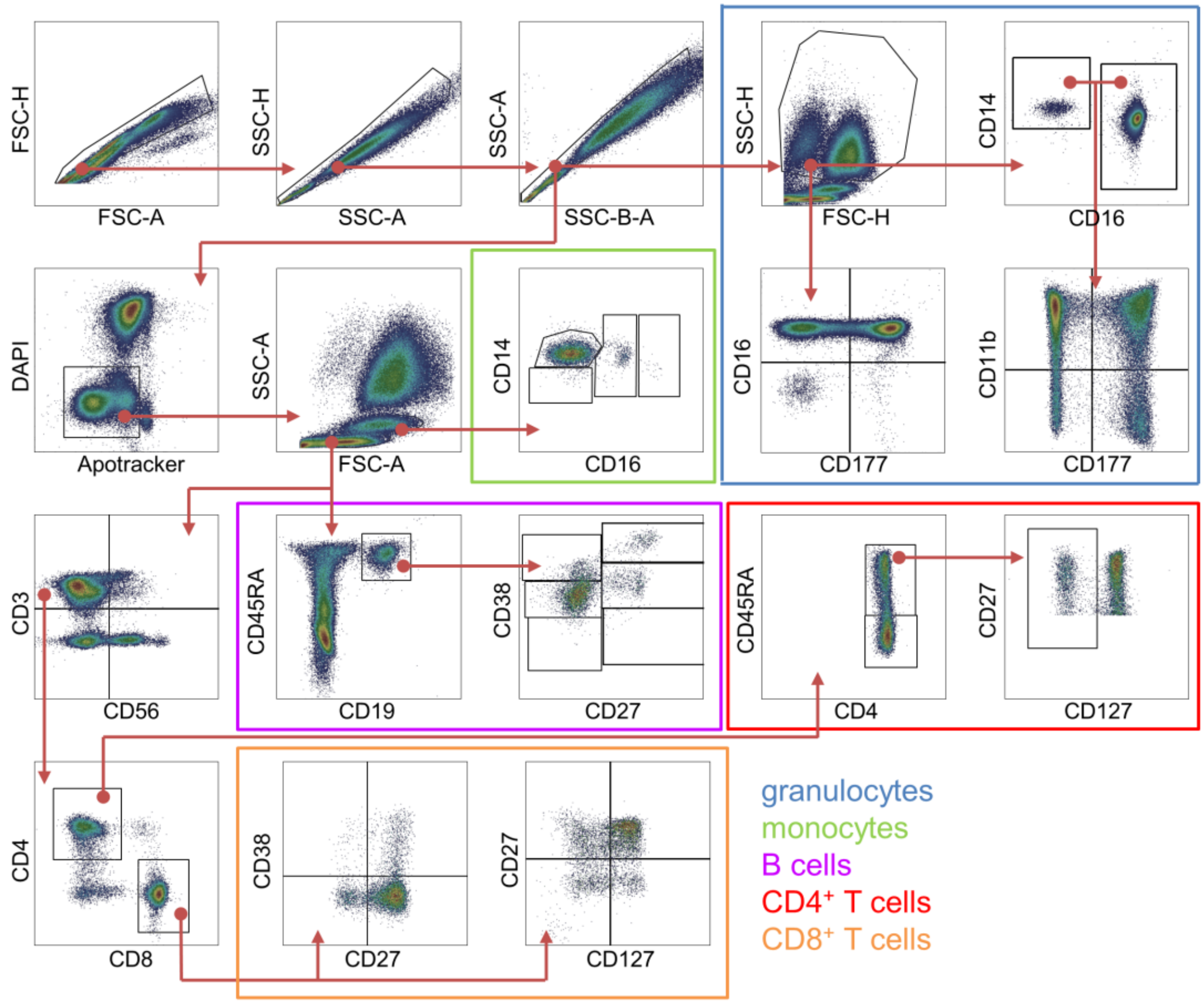


**Supplementary Fig. 6 Gating scheme for the 24-colour immune-phenotyping of peripheral whole blood cells.** Arrows indicate the hierarchical gating steps.

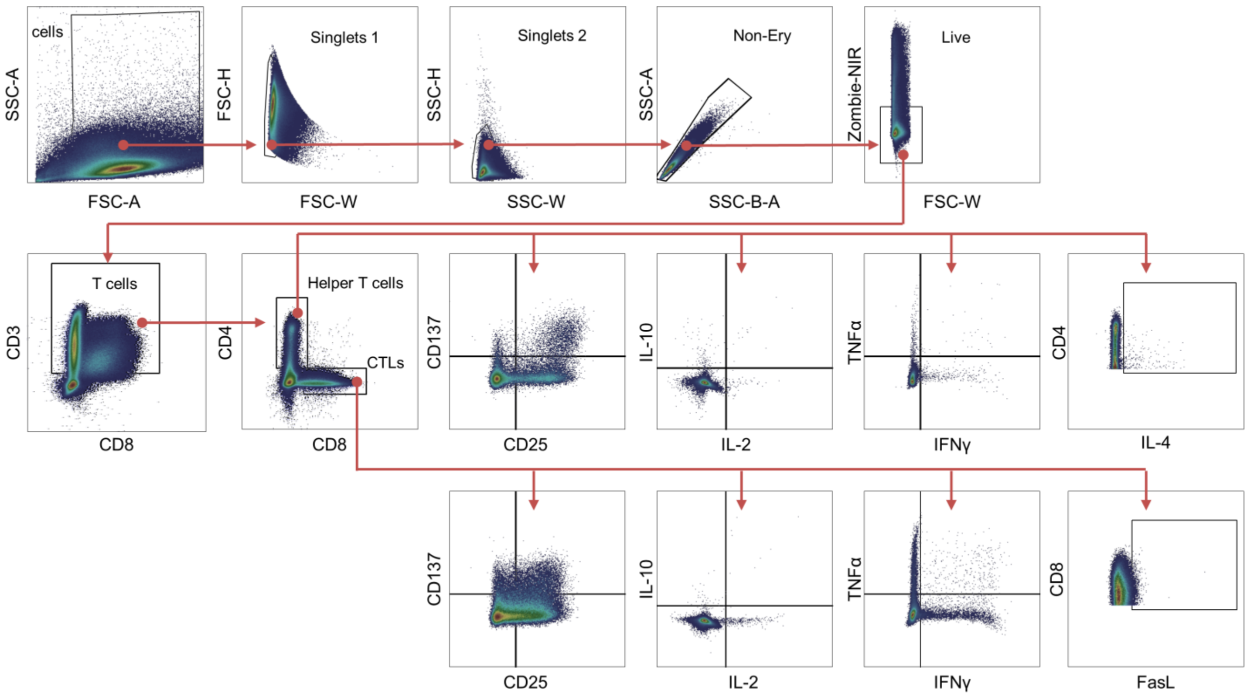


**Supplementary Fig. 7 Gating scheme for the intracellular cytokine staining assay, here exemplified for PMA/Ionomycin stimulated PBMC.** Expression of the activation markers CD25, CD137 as well as the cytokines IL-2, IL-10, IFNγ and TNFα were analyzed on $CD4^+$ and $CD8^+$ cells, respectively. IL-4 production was analyzed only on $CD4^+$ cells while FasL expression was examined for $CD8^+$ cells only. Arrows indicate the hierarchical gating steps.

**Supplementary Table 1 Immune status during early AZD1222 vaccination response**

| | Day 0 (n=18) cells/μL [median] (IQR) | Day 2 (n=15) cells / μL [median] (IQR) | Day 6 (n=17) cells / μL [median] (IQR) | Day 13 (n=16) cells / μL [median] (IQR) | Day 20 (n=16) cells / μL [median] (IQR) | p-value (Kruskal-Wallis) |
|---|---|---|---|---|---|---|
| granulocytes | 2394 (1842-3093) | 632 (479-1101) | 2004 (1618-2232) | 2086 (1427-2862) | 2107 (1355-2850) | < 0.0001 |
| monocytes | 369 (253-448) | 269 (226-311) | 278 (252-412) | 252 (189-349) | 297 (174-372) | 0.3110 |
| lymphocytes | 1846 (1788-2658) | 1014 (698-1262) | 1803 (1560-2689) | 1925 (1585-2687) | 1827 (1439-2338) | 0.0009 |
| B cells | 216 (140-287) | 79 (57-160) | 162 (126-261) | 246 (187-409) | 151 (122-278) | 0.0019 |
| T cells | 1102 (873-1524) | 560 (416-695) | 1114 (969-1419) | 1178 (893-1416) | 1005 (750-1230) | 0.0004 |
| CD4+ T cells | 684 (521-1092) | 333 (262-466) | 734 (596-936) | 799 (584-954) | 637 (494-822) | 0.0026 |
| CD8+ T cells | 325 (248-358) | 149 (89-185) | 299 (252-337) | 291 (214-351) | 252 (209-313) | < 0.0001 |

**Supplementary Table 2 Immune status during early BNT162b2 vaccination response**

| | Day 0 (n=18) cells/μL [median] (IQR) | Day 2 (n=17) cells / μL [median] (IQR) | Day 6 (n=18) cells / μL [median] (IQR) | Day 13 (n=16) cells / μL [median] (IQR) | Day 20 (n=17) cells / μL [median] (IQR) | p-value (Kruskal-Wallis#) (one-way ANOVA*) |
|---|---|---|---|---|---|---|
| granulocytes | 1330 (1132-2100) | 1240 (1043-1683) | 1571 (1239-2246) | 1173 (831-1836) | 1646 (1098-2221) | 0.3834# |
| monocytes | 345 (215-420) | 329 (251-391) | 303 (253-371) | 331 (257-389) | 307 (216-407) | 0.9404* |
| lymphocytes | 1829 (1332-2076) | 1816 (1335-2023) | 1756 (1603-2097) | 1746 (1441-2674) | 1704 (1492-2401) | 0.7708* |
| B cells | 158 (125-260) | 147 (132-210) | 233 (136-388) | 178 (120-292) | 229 (125-307) | 0.0885* |
| T cells | 945 (726-1194) | 799 (705-1104) | 946 (641-1053) | 986 (648-1219) | 1046 (761-1409) | 0.4438# |
| CD4+ T cells | 558 (490-617) | 511 (409-578) | 543 (370-606) | 589 (386-754) | 623 (513-849) | 0.3272# |
| CD8+ T cells | 249 (183-381) | 236 (167-345) | 241 (197-361) | 256 (196-424) | 302 (230-457) | 0.6468# |

**Supplementary Table 3 Responders to vaccines**

| | IgM [% responders] | | IgG [% responders] | | IgA [% responders] | |
|---|---|---|---|---|---|---|
| vaccine | AZD1222 | BNT162b2 | AZD1222 | BNT162b2 | AZD1222 | BNT162b2 |
| day 0 | 0 | 0 | 5.3 | 0 | 0 | 0 |
| day 2 | 0 | 0 | 0 | 0 | 0 | 0 |
| day 6 | 0 | 0 | 11.1 | 0 | 0 | 5.6 |
| day 13 | 52.9 | 26.7 | 47.1 | 86.7 | 11.8 | 100 |
| day 20 | 52.9 | 47.1 | 82.4 | 100 | 23.5 | 100 |
| Fisher´s exact test [p value] | 0.033 | | 0.55 | | 0.040 | |
